# Trend Analysis of Hepatitis B and C Virus Infections among Patients at Ambo General Hospital, West Shoa, Ethiopia: Retrospective Study

**DOI:** 10.1101/2024.10.31.24316472

**Authors:** Mulugeta Getachew, Amanuel Teferi, Edosa Kebede

## Abstract

**Background:** Hepatitis B and C viruses affect the liver and can cause wider range of disease outcomes. Chronic HBV and/or HCV infection leaves a person susceptible to major liver diseases such liver cirrhosis or hepatic cell cancer later in life. They bear the greater portion of the mortality and morbidity associated with hepatocellular carcinomas and liver cirrhosis.

**Methods:** A retrospective laboratory record review was conducted at Ambo General Hospital from January 2018 to December 2021. The retrieved data included the date of examination, age, sex and laboratory results of the HBV and HCV. Data were summarized and presented in the form of tables, figures, and frequencies to present the results. The data were analyzed using SPSS and Microsoft Excel.

**Results:** Over the course of 4 years, a total of 5675 individuals were examined for hepatitis at Ambo general hospital. Of the total individuals examined, 365(6.4%) and 15(2.1%) were positive for HBV and HCV respectively. HBV and HCV mixed infections accounted for 0.6% of the cases. HBV was highest prevalent in males (12.1%) than females (5.72%), while among seropositive of HCV, 7(5.7%) were males and 4(1.0%) were females. HBV and HCV were more prevalent among individuals aged from 45–54 years old 11(11.6%) and 35–44 years old 4(5.6%) respectively. A high percentage of HBV (7.1%) and HCV (5.5%) were seen in the years of 2018 and 2020 respectively.

**Conclusion:** According to WHO criteria, the total prevalence of hepatitis B and C in our study is moderate, and the prevalence of HBV is significantly higher than that of HCV. Although, there are annual fluctuations in the prevalence. As a result, it is critical to improve coverage of services in healthcare facilities and raise community awareness regarding the means of transmission, prevention and control of hepatitis B and C virus infection.

## Introduction

Hepatitis is an inflammation of the liver that can be caused by infectious and non-infectious agents such as viruses, bacteria, fungi, parasites, alcohol, drugs, self-immune diseases, and metabolic diseases. The significant causes of hepatitis are viruses; such as hepatitis A, B, C, D and E viruses. The most frequent causes of viral hepatitis among them are the hepatitis B virus (HBV) and the hepatitis C virus (HCV). Both HBV and HCV infections are extremely contagious and can spread vertically, through sexual contact, and through blood transfusions[1] .

Viral hepatitis is a serious health problem globally and cause acute and chronic hepatitis which can result in liver damage (cirrhosis), liver failure, liver cancer and even death. Viral hepatitis causes over 1.4 million deaths annually, with HBV and HCV accounting for roughly 96% of these cases[2]. There were 257 and 10 million HBV and HCV infections worldwide, respectively. Of these, 80 million have active viremia infections [6, 7]. If left untreated, up to 30% of chronic HBV and 20% of chronic HCV cases develop liver cirrhosis and hepatocellular carcinoma, which are the sixth most common cancer and the third cause of cancer-associated deaths worldwide[3]. The prevalence of HBV infection varied from high (≥8%) to intermediate (2–7%) and low (3.5%), moderate (1.5–3.5%), and low (<2%). Likewise, HCV infection is high (>3.5%), moderate (1.5– 3.5%), and low (<1.5%) prevalence according to the World health organization (WHO)[4].

While the prevalence of HBV and HCV infection is distributed globally, the great majority of those who are affected live in low- and middle-income nations [5]. In Africa, the prevalence of HBV is 6·1%, with about 87,890 deaths annually. However, the Sero-prevalence differs depending on sex, ethnicity, and rural residence[6], [7].

In Ethiopia, there are various studies indicated the trends of HBV and HCV in different areas. They showed viral hepatitis remains a public health problem. A systematic review and meta-analysis clinical study showed that liver disease accounted for 12% hospital admissions and 3% hospital mortality due to HBV and HCV infections[8]. In the study area, there is no adequate data indicating the progress of HBV and HCV infections. Therefore, the aim of this study was to determine a trend of Hepatitis B and C virus infections among patients who have been tested at Ambo General Hospital from 2018-2021.

## Materials and Methods

### Study design and setting

A facility-based retrospective study was conducted from April to July 2022 among patients who have been tested for hepatitis B and C viruses at Ambo General Hospital from January 2018 to December 2021. Based on census of 2021 population of Ambo city has 64684 people. There are two public hospitals in Ambo city; Ambo university referral Hospital and Ambo General Hospital.

### Source and study population

All HBV and HCV suspected patients at Ambo General Hospital from January 2018 to December 2021 were the source population. Whereas, all patients who have been tested for HBV and HCV and whose results recorded on laboratory registration log book at Ambo General Hospital from January 2018 to December 2021 within the specified period were study population to this study.

## INCLUSION AND EXCLUSION CRITERIA

### Inclusion Criteria

All HBV and/or HCV suspected and screened patients, whose results recorded on laboratory registration logbook from January 2018 to December 2021 and fulfilled all requested format information like age, sex, date, month, years of examination and hepatitis status were included in this study.

### Exclusion Criteria

Patient records without full information were excluded

## DATA COLLECTION PROCEDURE

Three medical laboratory technologists extracted data for all HBV and/or HCV suspected and tested patients whose findings were documented on the laboratory registration logbook from January 2018 to December 2021. Age, sex, the year of screening, and the HBV and HCV status were gathered as pertinent characteristics.

### Statistical analysis

Using SPSS version 25.0 we conducted univariate and multivariate regression to estimate odds ratios with 95% confidence intervals (CI) for potential predictors for HBV and HCV infection. We included demographic variables such as sex, and age. Microsoft Excel was used to evaluate the trend of hepatitis.

## ETHICAL CONSIDERATION

All procedures performed in this study were performed in accordance with the guideline and regulations approved by the Research Ethics Review Committee of the Ambo University, College of Health Sciences and Referral hospital, Department of medical laboratory Sciences with a reference number to get permission for collecting the data.

## Results

### Socio-demographic characteristics

A total of 5,675 serologically tested patients were included in our study. Of these, 637 (11.2%) were male and 5,038 (88.8%) were females, so the majority of study participants were females. The majority of study subjects were aged from 15–34 years old 4,891 (86.2%) [Table 1].

**Table 1:**
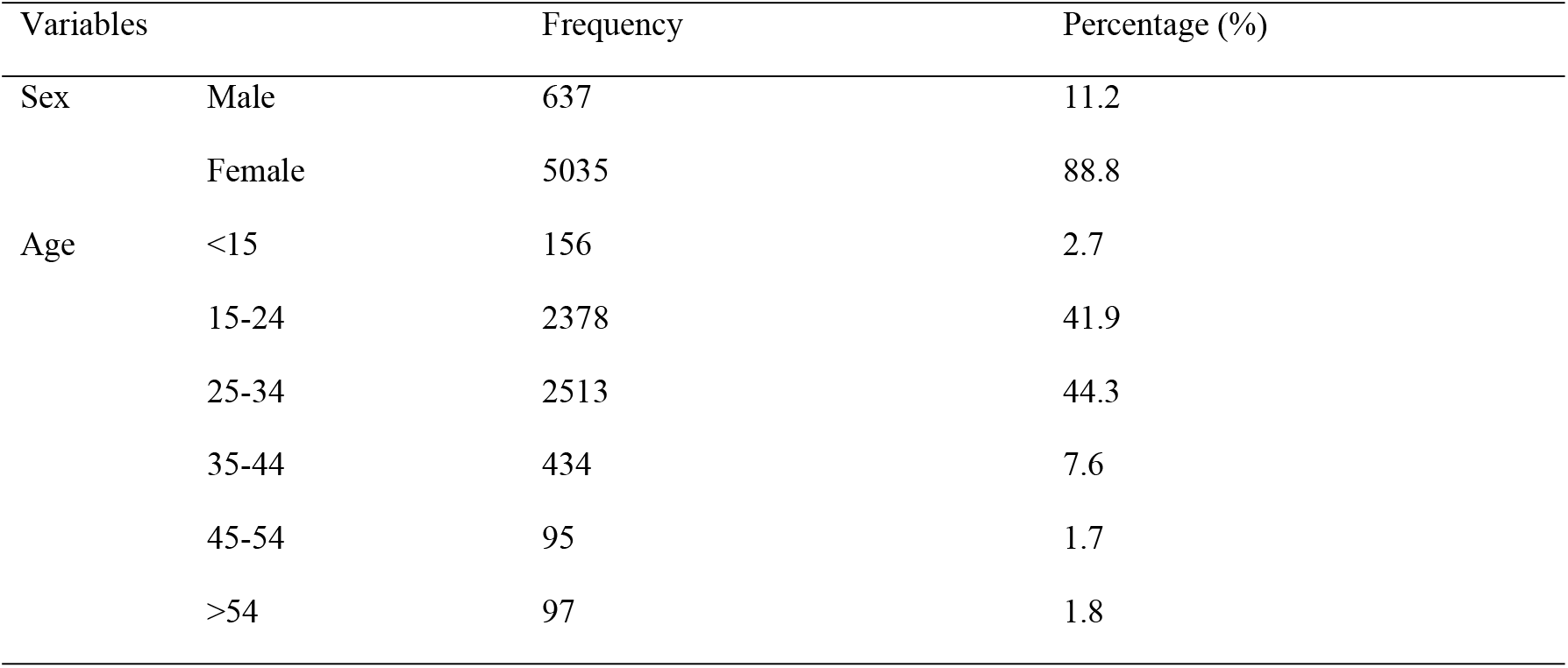
Socio-demographic characteristics among people who were tested for HBV and HCV from January 2018 to December 2021 at Ambo General Hospital.

### Seroprevalence of HBV and HCV

The overall prevalence of HBV and HCV was 365/5674(6.4%) and 11/524(2.1%) respectively. Among the study participants, the prevalence of HBV was 77(12.1%) among males and 288(5.72%) among females, while the positivity of HCV were 7/122(5.7%) among males and 4/402(1.0 %) among females. Hepatitis B virus was predominantly higher among individuals aged 45-54 years old (11.6%) followed by >54 years old (9.28%). On the other hand, the prevalence of hepatitis C virus was high among the age group of 35–44 years old (5.6%) followed by 45-54 years old (4.0%). Moreover, a high percentage of HBV (7.1%) and HCV (5.5%) were seen in the years of 2018 and 2020 respectively [Table 2].

**Table 2:**
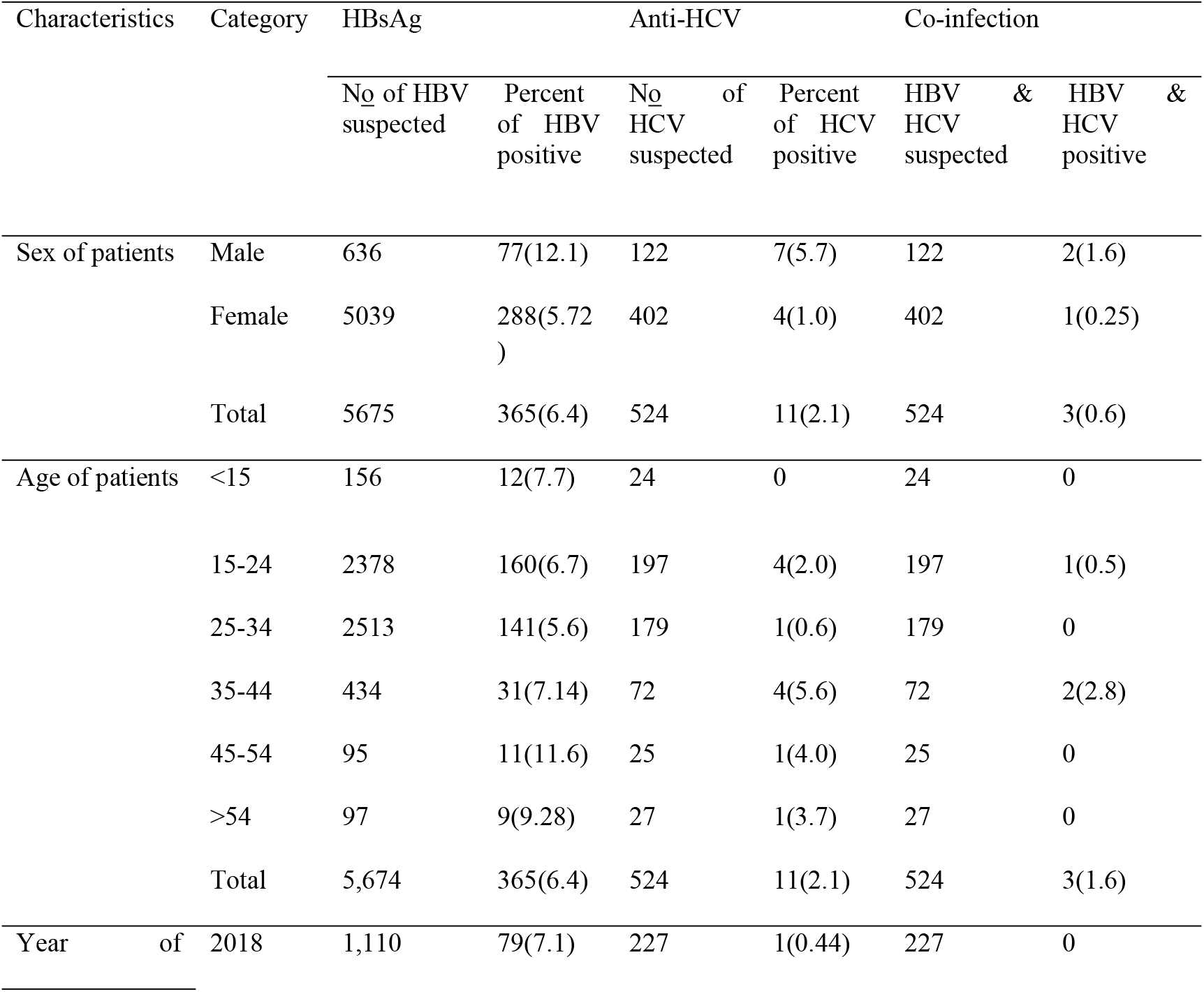
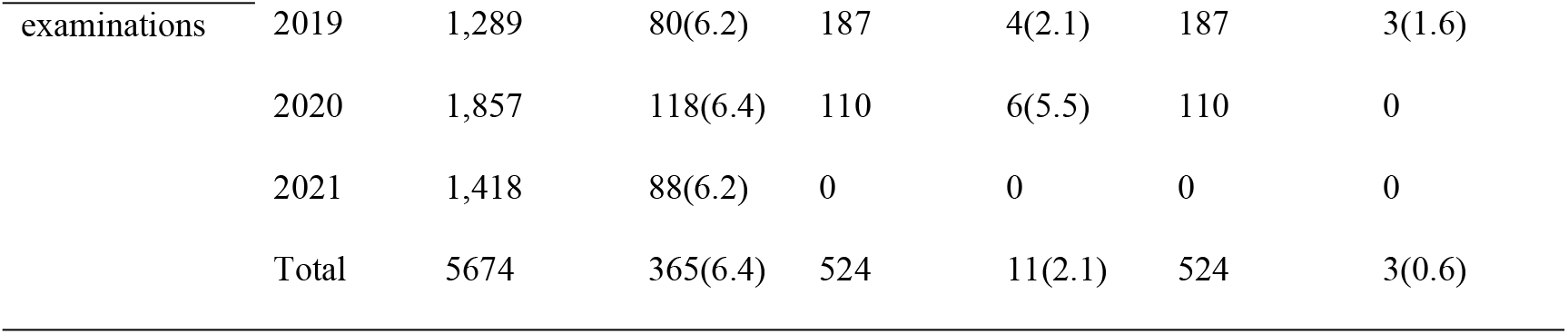
Prevalence of HBV and HCV infection among people who were tested for HBV and HCV from January 2018 to December 2021 at Ambo General Hospital.

### Factors associated with hepatitis B virus infection

According to the bivariate analysis, both sex and age variables showed a P-value less than 0.25 and transported to multivariate analysis. Accordingly, in multivariate analysis, being a male was significantly associated with HBV infection [AOR = 2.186 (95% CI: 1.652, 2.892, p <0.001] and age groups not showed significant association with HBV infection [Table3].

**Table 3:** Factors associated with hepatitis B among study participants at Ambo General Hospital from January 2018 to 2021December.

| Variables |  | HBV |  | COR(95% CI) | P-value | AOR(95% CI) | p-value |
| --- | --- | --- | --- | --- | --- | --- | --- |
|  |  | Pos (%) | Neg (%) |  |  |  |  |
| Sex | Male | 12.1 | 87.9 | 2.272 (1.741, 2.966) | <0.001 | 2.186 (1.652, 2.892) | <0.001 |
|  | Female | 5.72 | 94.28 | 1 |  | 1 |  |
| Age | <15 | 7.7 | 92.3 | 1 | 1 | 1 |  |
|  | 15-24 | 6.8 | 93.2 | 1.213 (.492, 2.996) | .675 | .989 (.398, 2.460) | .981 |
|  | 25-34 | 5.6 | 94.4 | 1.402 (.693, 2.834) | .347 | 1.033 (.502, 2.128) | .929 |
|  | 35-44 | 7.14 | 92.8 | 1.701 (.839, 3.447) | .140 | .875 (.423, 1.810) | .718 |
|  | 45-54 | 11.6 | 88.4 | 1.315 (.605, 2.859) | .490 | 1.028 (.467, 2.263) | .945 |
|  | >54 | 9.28 | 90.7 | .772 (.305, 1.957) | .586 | 1.368 (.536, 3.490) | .512 |

### Factors associated with hepatitis C virus infection

Bivariate logistic regression analysis were performed to assess the association between dependent and independent variables. According to the bivariate analysis, only sex of the study participants showed a P-value less than 0.25. However, being a male was not significantly associated with HCV infection [AOR = 2.262(95% CI: .797, 6.420, p = 0.125] [Table 4].

**Table 4:** Factors associated with hepatitis C among study participants at Ambo General Hospital from January 2018 to 2021December.

| Variables |  | HCV |  | COR(95% CI) | P-value | AOR(95% CI) | p-value |
| --- | --- | --- | --- | --- | --- | --- | --- |
|  |  | Pos (%) | Neg (%) |  |  |  |  |
| Sex | Male | 7.5 | 92.5 | .442 (.156, 1.254) | 0.125 | 2.262(.797, 6.420) | 0.125 |
|  | Female | 3.5 | 96.5 | 1 | 1 | 1 |  |
| Age | <15 | 6.25 | 93.75 | 1 | 1 |  |  |
|  | 15-24 | 4.8 | 95.2 | 1.333 (.146, 12.210) | .799 |  |  |
|  | 25-34 | 3.5 | 96.5 | 1.844 (.180, 18.938) | .606 |  |  |
|  | 35-44 | 7.84 | 92.16 | .783 (.081, 7.560) | .833 |  |  |
|  | 45-54 | 4.2 | 95.8 | 1.533 (.089, 26.431) | .769 |  |  |
|  | >54 | 4.00 | 96 | 1.600 (.093, 27.547) | .746 |  |  |

### Trend prevalence of HBV and HCV infections

The trend prevalence of HBV and HCV was relatively fluctuating from year to year. The prevalence of HBsAg was 79/1110(7.1%) in 2018, 80/1289(6.2%) in 2019, 118(6.4%) in 2020 and 88(6.2%) in 2021. The anti-HCV prevalence was 1/227(0.44%) in 2018, 4/187(2.1%) in 2019, 6/110 (5.5%) in 2020 and in 2021 the test was not done [fig1 and fig 2].

**Fig 1:**
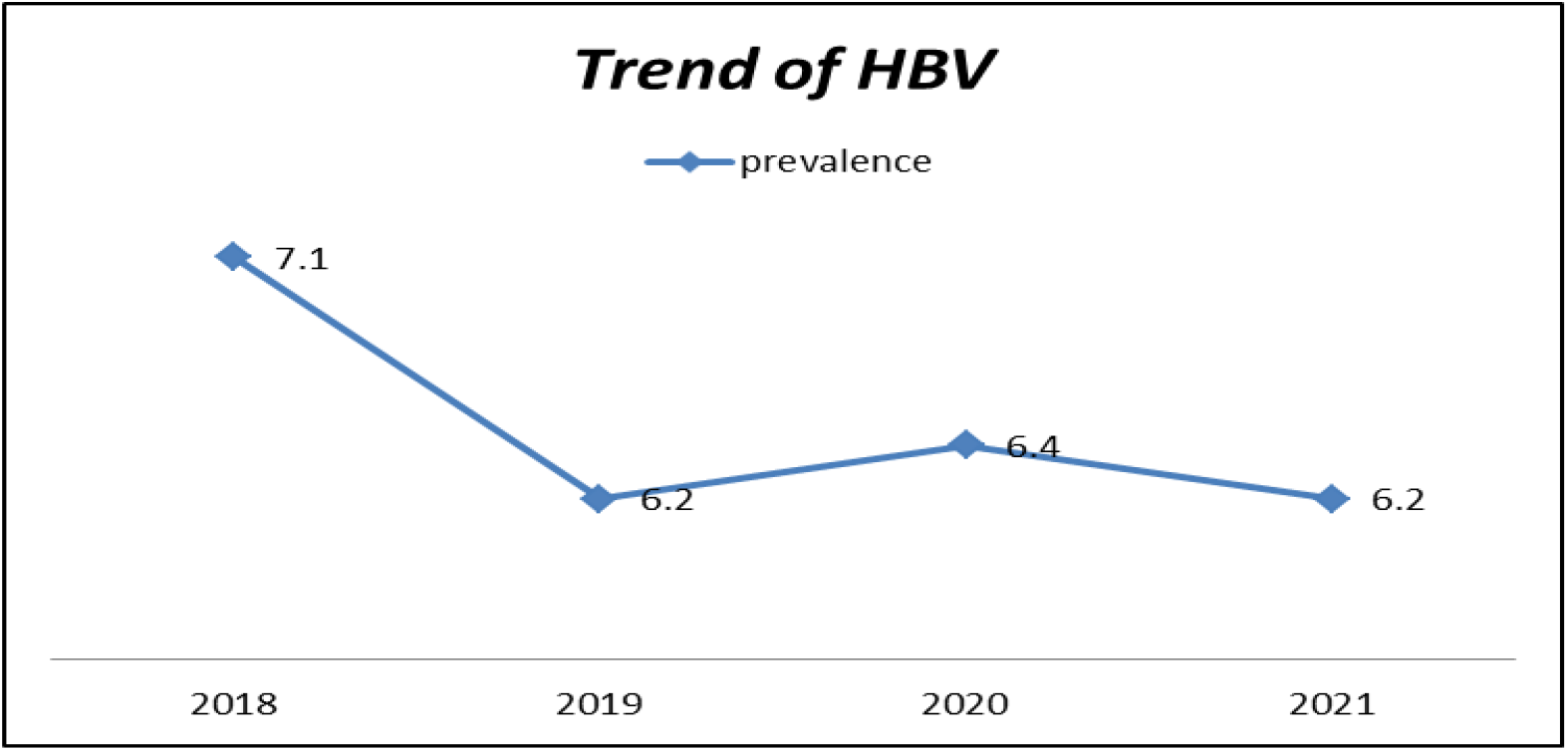
Trend prevalence of Hepatitis B virus

**Fig 2:**
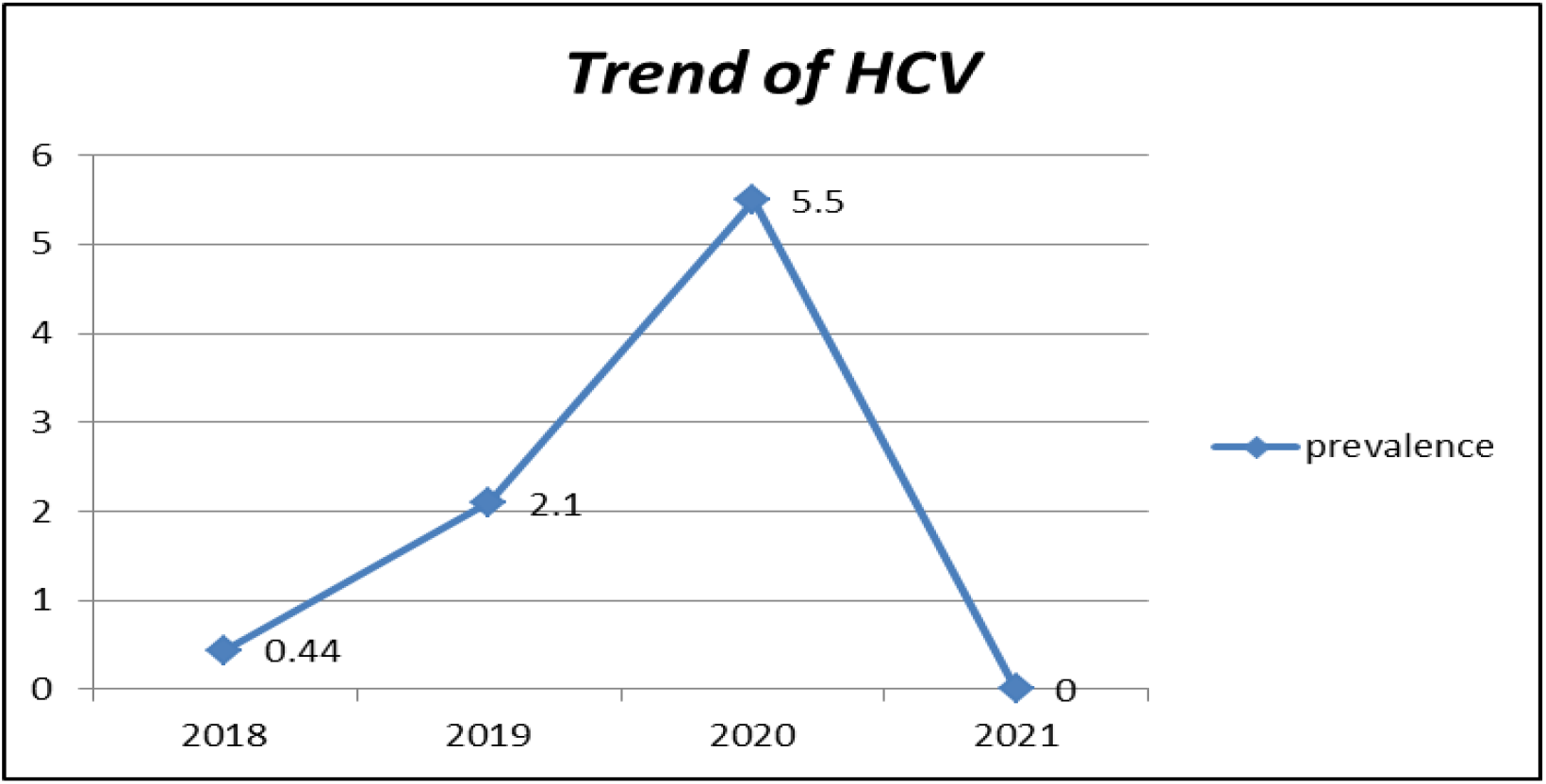
Trend prevalence of Hepatitis C virus

## DISCUSSION

In this study, the prevalence of HBV and HCV was 6.4% and 2.1% respectively. This prevalence is moderate according to the WHO[4]. The prevalence of HBsAg is similar with study conducted among women attending antenatal clinics in West Hararghe public hospitals, Oromia region, Ethiopia that was 6.1%[9]. Again, our study also relatively similar with study conducted among migrants attended at a center in Madrid in Spain that was 6% for HBV and it showed similar prevalence of HCV (2%)[10].

However, the prevalence of HBV (6.4%) in this study is relatively higher than the prevalence HBV(3.9%) reported from a recent retrospective study done in Addis Alem Primary Hospital, Bahir Dar, Northwest Ethiopia[11] and study conducted in Central West Argentina(1.8%)[12]. This result variation might be due to the differences in sociocultural. On the other hand, the prevalence of this study relatively lower than the study conducted among clients visiting ‘Tefera Hailu’ memorial hospital, Sekota, Northern Ethiopia, with a prevalence of 21.16 % for HBV [13] and the study conducted among patients attending Gondar University Hospital with a prevalence of 14.4% for HBV and 12.4% for HCV[14]. This difference might be due to sample size and cultural variations.

The overall prevalence of HBV and HCV co-infection was 0.6%. This finding is lower than with the study reported in Ethiopia, Gondar 2%[15]. This is due to the difference in the sample size of the study participants and socio-cultural of the society. The result of our study showed that most of the positive study participants were found in the age category of 45-54 years old (11.6%) for HBV and 35-44 (5.6%) for HCV. This result is not similar to the result of the study conducted at Gondar University that showed HBV was found to be more prevalent among study subjects aged 25–34 years old (30.2%) and HCV was more prevalent among those aged 15–24 years old (30.7%)[14]. In our study, the prevalence of hepatitis was more prevalent among males than females, which was similar with a study conducted in Sierra lione [16]. However, our result was not similar with study conducted in Gondar University Hospital[14]. This might be due to the majority of our study participants were females. The trend both of HBV and HCV was relatively fluctuating from year to year. The highest prevalence was recorded in 2018(7.1%) for HBV and in 2020 (5.5%) for HCV.

## CONCLUSION

The prevalence of HBV in our study area is considerably higher than HCV among patients who have tested at the hospital. The prevalence of both HBV and HCV was high in males than females. HBV more prevalent in age grouped from 45-54 while HCV more prevalent in age grouped from 35-44. These groups of age are working age, so it affects the economy of the countries. The prevalence of both HBV and HCV was fluctuating from year to year. This indicates that follow up should be undertaken by providing the materials necessary for the diagnosis that can prevent the underestimation of the prevalence. Therefore, consideration should be given by the government and non-government organizations on diagnosis, prevention & control of this virus transmission in the study area.

## Data Availability

The data that support the findings of this study are available upon reasonable request from the corresponding author

## Funding sources

There was no funding to conduct the research.

## Acknowledgments

The authors would like to extend their gratitude to Ambo General Hospital for their willingness and cooperation during the data collection.

## Author Contributions

Mulugeta Getachew, Amanuel Teferi, and Edosa Kebede are involved in the conception of the research idea, design and data collection, analysis and interpretation of the finding. The manuscript was written by Mulugeta Getachew, and all the other authors have read and approved the final manuscript.

### Author Biographies

Mulugeta Getachew has MSc in Medical Parasitology from Jimma University and BSc in Medical Laboratory Science from Hawassa University. His research interest is public health, particularly with infectious disease and non-infectious diseases affecting communities. He is serving as lecturer at the college of Health Sciences and Referral Hospital, Ambo University.

Amanuel Teferi has MSc in Medical Microbiology from Addis Ababa University and BSc in Medical Laboratory Science from Hawassa University. His research interest is public health, particularly with infectious disease and non-infectious diseases affecting communities. He is serving as lecturer and department head of Medical Laboratory Science at the college of Health Sciences and Referral Hospital, Ambo University.

Edosa Kebede has MSc in Medical Microbiology from Jimma University and BSc in Medical Laboratory Science from Jimma University. His research interest is public health, particularly with infectious disease and non-infectious diseases affecting communities. He is serving as lecturer and participating in different committees at the department level and college of Health Sciences and Referral Hospital, Ambo University.

## Notes

### Competing Interest Statement

The authors have declared no competing interest.

### Funding Statement

The author(s) received no specific funding for this work.

